# Association between the Mediterranean diet and prostate cancer risk in a Greek population

**DOI:** 10.1101/2020.08.13.20172999

**Authors:** Alexandros Vaioulis, Kiouvrekis Yiannis, Konstantinos Perivoliotis, Gravas Stavros, Tzortzis Vasilios, Karatzas Anastasios

**Affiliations:** Faculty of Medicine, School of Health Sciences, University of Thessaly, Department of Urology; University of Nicosia, Nicosia, Cyprus; University of West Attica, Department of Biomedical Sciences, Athens, Greece; Faculty of Medicine, School of Health Sciences, University of Thessaly, Department of Surgery

**Author notes:** To whom correspondence should be addressed: **Anastasios D. Karatzas**, MD, PhD, FEBU, FEAPU, Urology Department, Faculty of Medicine, School of Health Sciences, University of Thessaly, Biopolis, 41110, Larissa, Greece.

**Keywords:** Mediterranean diet, prostate, cancer, biopsy, PSA

## Abstract

**Purpose:** Dietary modifications have been correlated with survival in several neoplasia, such as prostate cancer. The present study was designed to investigate the association between the Mediterranean diet and prostate cancer risk.

**Methods:** A retrospective analysis of patients with a high suspicion of suffering from prostate cancer (PCa) who underwent prostate biopsy was performed. According to histopathology, two groups were generated, the PCa group and the Healthy group. The dietary profile of our study population was implemented, based on a modified MDS (Mediterranean Diet Score) questionnaire. A logistic regression model was used for the interpretation of our data.

**Results:** In total, 431 patients with prostate cancer and 279 healthy men were included in our study. The demographic characteristics of the patients were comparable. Daily consumption of white meat (OR: 0.59), dairy products (OR: 0.64), nuts (OR: 0.63) and whole grains (OR: 0.55) was higher in healthy males. Infrequent consumption of vegetables was linked with an increased rate of PCa (OR: 2.55). Interestingly daily consumption of processed meat rates was higher in healthy men. However, a significant correlation between specific intake products or frequency and the incidence of PCa was not established.

**Conclusions:** Although, an association between the dietary patterns and PCa was not determined, components consumption patterns displayed a higher daily intake rate of white meat, dairy products, nuts and whole grains. Further prospective trials are required to validate the effect of Mediterranean diet in the incidence and mortality of PCa patients.

## INTRODUCTION

Prostate cancer (PCa) is the second most common malignancy in men, with almost 1 million newly diagnosed cases every year (1). It represents the 14% of primary diagnosed tumors and 6% of cancer related mortality in male population. An increasing trend is documented, with PCa reaching 1.7 million new cases and 0.5 million deaths by 2030 (2).

Due to the high prevalence and the derived socioeconomic costs, various factors have been evaluated for their potential prognostic role in PCa survival. These include demographic characteristics (e.g. age, gender, nationality, heredity), clinical features (e.g. Gleason score, prostatic specific antigen level, tumor stage, resection margins), molecular biomarkers (e.g. Ki67, epidermal growth factor, vascular endothelial growth factor, p53) and dietary habits (3).

Habitational motives and dietary patterns have been identified as survival outcome modifying factors in various malignancies (4). Taking into consideration the lower overall cancer incidence in Mediterranean countries compared to USA, the value of the Mediterranean diet (MD) has been thoroughly investigated. In a randomized controlled trial of 7447 participants, a negative correlation between the consumption of specific Mediterranean nutrients and cancer mortality was identified (5). Furthermore, recent pooled analyses confirmed the inverse association between the adherence to Mediterranean diet and cancer risk, mortality and recurrence rates (4, 6).

Various studies attempted to associate the Mediterranean diet with PCa survival. Trichopoulou et al. estimated that compliance with the MD standards would result in a 10% reduction of PCa incidence in USA alone (7). Although, Kenfield et al. in a large retrospective analysis failed to link the Mediterranean diet with advanced PCa stage or disease progress, their results confirmed a lower mortality rate in non-metastatic PCa patients who followed MD(8). Bosire et al., however, could not identify any significant correlation between PCa outcomes and Mediterranean diet (9).

In the latter body of evidence, specific nutritional components of the Mediterranean diet have been exclusively examined for their prognostic and preventive role. Olive oil, as the main source of fat acids in the Mediterranean diet, was not associated with the PCa incidence in various studies. Fruit and vegetables display an inhibitory effect in the PCa frequency, although results are conflicting. Despite the fact that cereals increase fiber intake, the analogous consumption of red and processed meat seem to positively correlate with PCa (10, 11). Interestingly, a meta-analysis of 13 studies suggested that the consumption of cruciferous vegetables significantly correlated with a reduced PCa risk (12).

As to the aforementioned, the present study was designed to estimate the possible association between the Mediterranean dietary profile and the prevalence of prostate cancer in a rural area of Greece.

## METHODS

### Study Protocol

Between 2003 to 2018, patients with a high suspicion of suffering from PCa entered our study in a retrospective manner. The eligibility criteria were prostatic specific antigen (PSA) level ≥4ng/ml, or a positive digital examination, or a lesion suspicious for malignancy in the transanal ultrasound examination (TRUS). The exclusion criteria included the introduction of a permanent urinary catheter, a recent urinary tract infection and the presence of a concurrent malignancy. In order to minimize bias, immigrant patients were not included in our protocol, since these populations display major epidemiological deviations due to environmental and dietary factors.

All patients underwent a prostate biopsy as previously described (13). According to histopathology, two groups were generated, the PCa Group (positive biopsies) and the Healthy Group (negative biopsies).

Informed consent was obtained from all individual participants included in the study. Prior to the onset of the present study, an approval from the Local Institutional Review Board was, also, received.

### Data Collection

Patient demographic data (age, BMI, PSA level, family history of PCa) were recorded. In order to estimate the dietary profile of the participants, the modified Mediterranean Diet (mMD) questionnaire, validated for the Greek population, was utilized (7,28). The mMD focuses on the intake of 10 items: fruit, red meat, white meat, roasted or fried meat, vegetables, fish, dairy products, cruciferous vegetables, nuts and whole grains. The consumption frequency for each item included 5 different types: ‘Never’, ‘<1 time per week’, ‘2-3 times per week’, ‘4-6 times per week’ and ‘Every day’.

### Endpoints

The primary endpoint of our study was to provide a descriptive consumption pattern overview of healthy and PCa individuals. Secondary endpoint was the investigation of a possible association between the intake of various components of Mediterranean diet and PCa incidence.

### Statistical Analysis

Prior to any evaluation, all data underwent Shapiro-Wilk test for normality. Independent samples t test was applied to continuous variables. For categorical variables, Pearson chi-square test was used. In case of a statistically significant result in the chi-square test, further subgroup comparisons were performed, through the calculations of Bonferroni adjusted z-tests. The Odds Ratios (OR) of significant comparisons were also reported. A logistic regression model was used for the identification of PCa incidence modifying dietary frequencies. Continuous data were reported as Mean (Standard Deviation), while categorical data were reported as N (Percentage). Statistical significance was considered at the level of P<0.05. All statistical analysis was conducted using IBM SPSS Statistics v23 software. This study is stated according to the STROBE guidelines (14).

## RESULTS

710 men were entered our study; 431 were diagnosed with PCa and 279 had negative TRUS biopsy results (Table 1). Mean age and BMI were 63.4(4.6) and 35.7(2.16), respectively. There was no statistically significant difference in terms of BMI, between the PCa and healthy study subgroups (38 vs 36.1, p=0.1). The mean PSA values of our sample were 7.21(1.8) and ranged from 4.2 to 15. In total, 59.2% of the PCa patients had a positive familiar history (p<0.001).

**Table 1:** Patient Characteristics

| Variable | Mean(SD) | Median(range) |
| --- | --- | --- |
| Age (years) | 63.4(4.6) | 62.2(52 – 80) |
| BMI | 35.7(2.16) | 33.5(22 – 35) |
| PSA (ng/dl) | 7.21(1.8) | 7.1(4.2 – 15) |
| Family history of PCa | 255/ 431 (59.2%) |  |

The mMD frequencies are reported in Table 2. A summary of these findings is displayed in Figure 1. Regarding fruit consumption, there was no overall significant difference between the two groups (Chi square p=0.132). The percentage of patients who never consumed or whose fruit intake rate ranged between less than 1 time to 4-6 times per week were comparable (p>0.05). However, a significant higher rate (p=0.02) of healthy individuals (33.6%) consumed fruit in a daily basis, compared to PCa patients (25.9%).

**Table 2:**
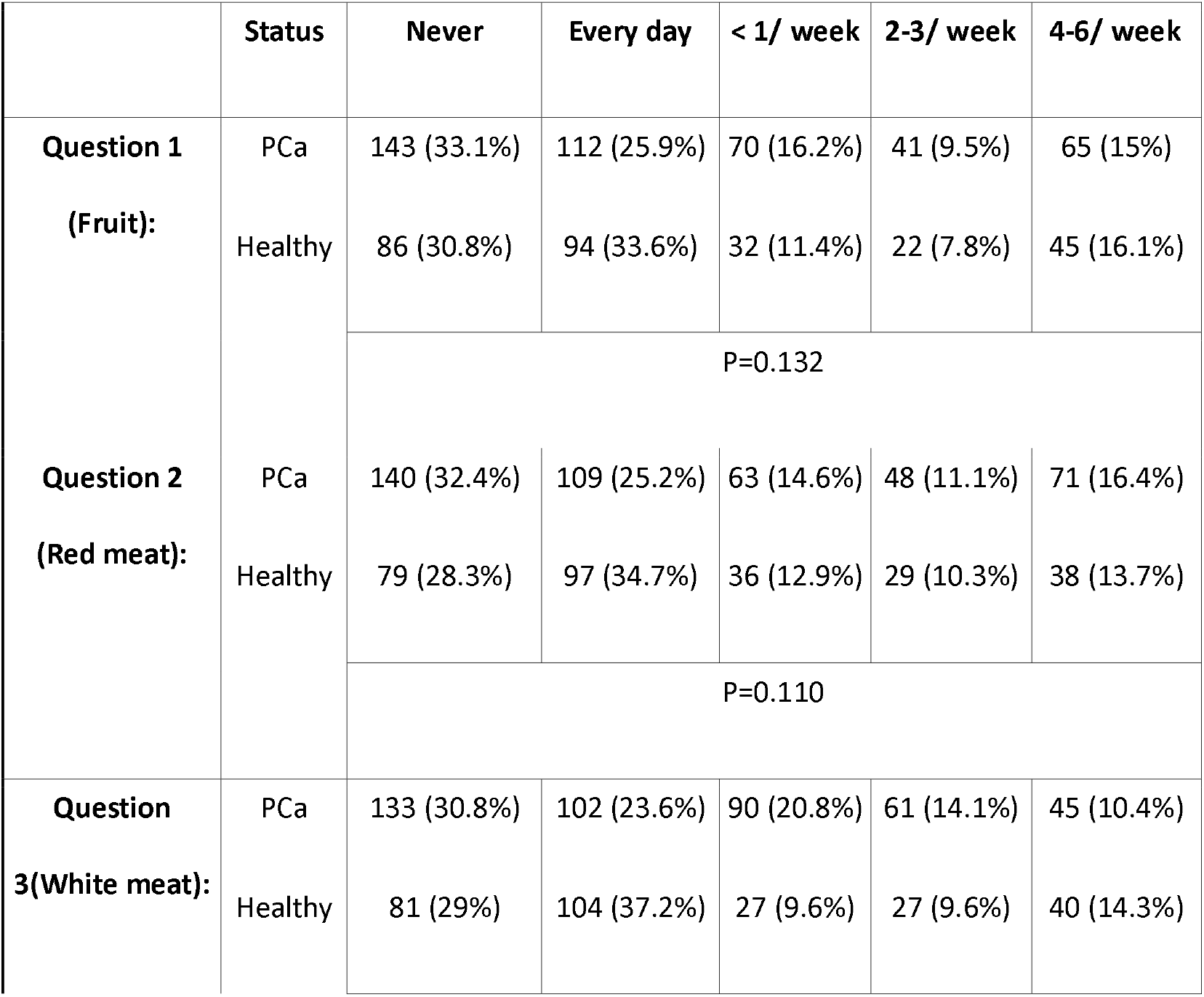
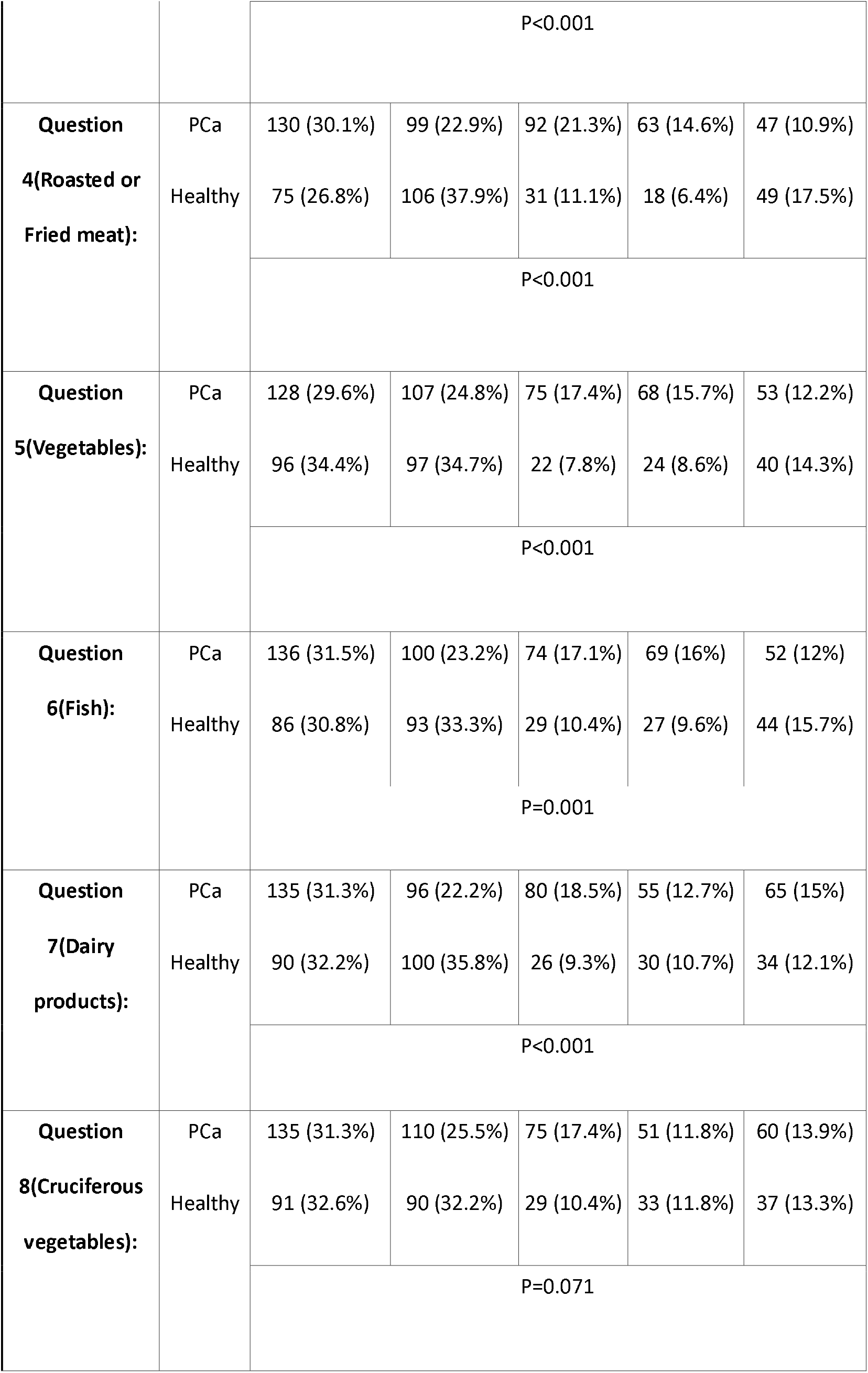
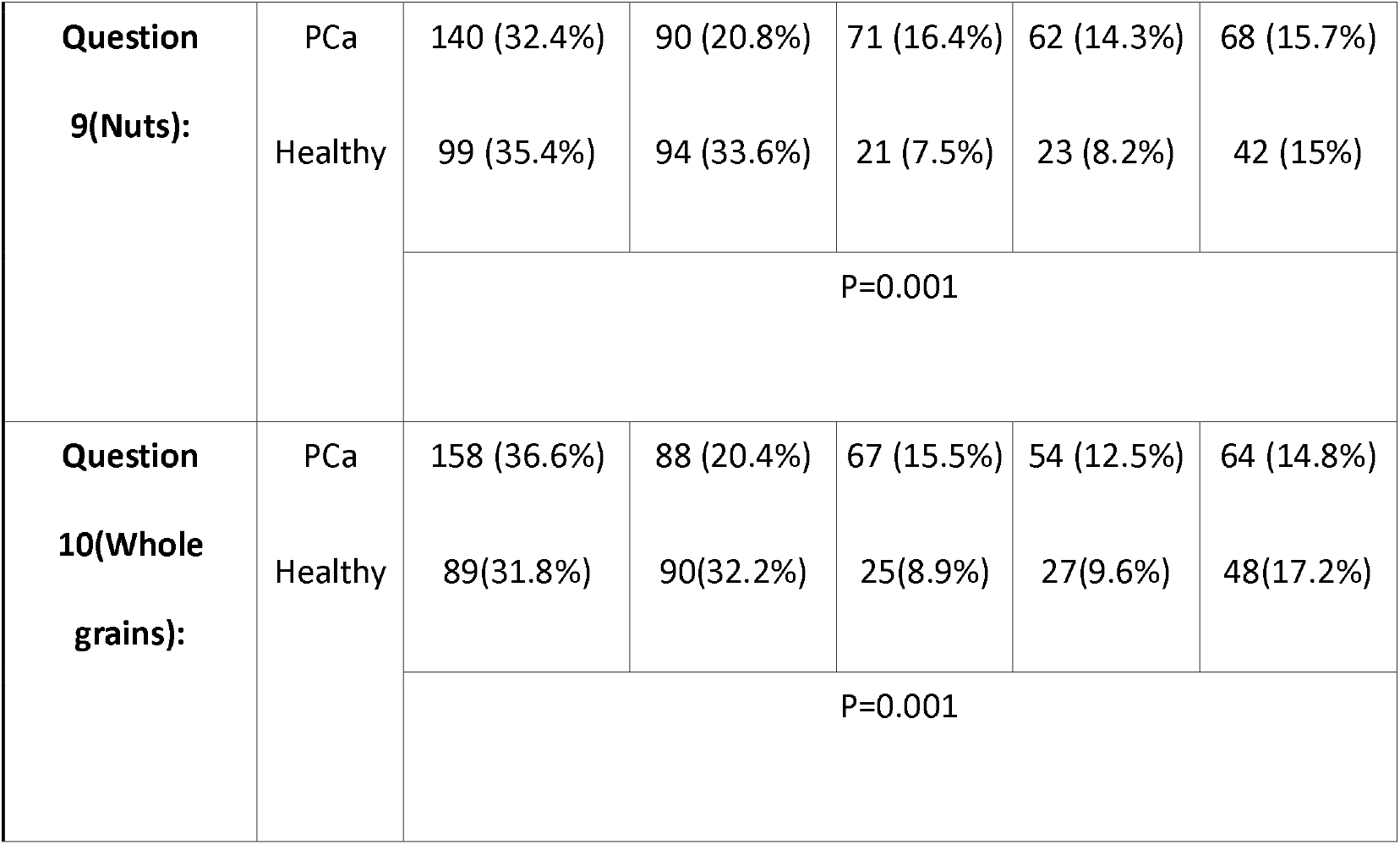
Questionnaire statistics

**Figure 1.**
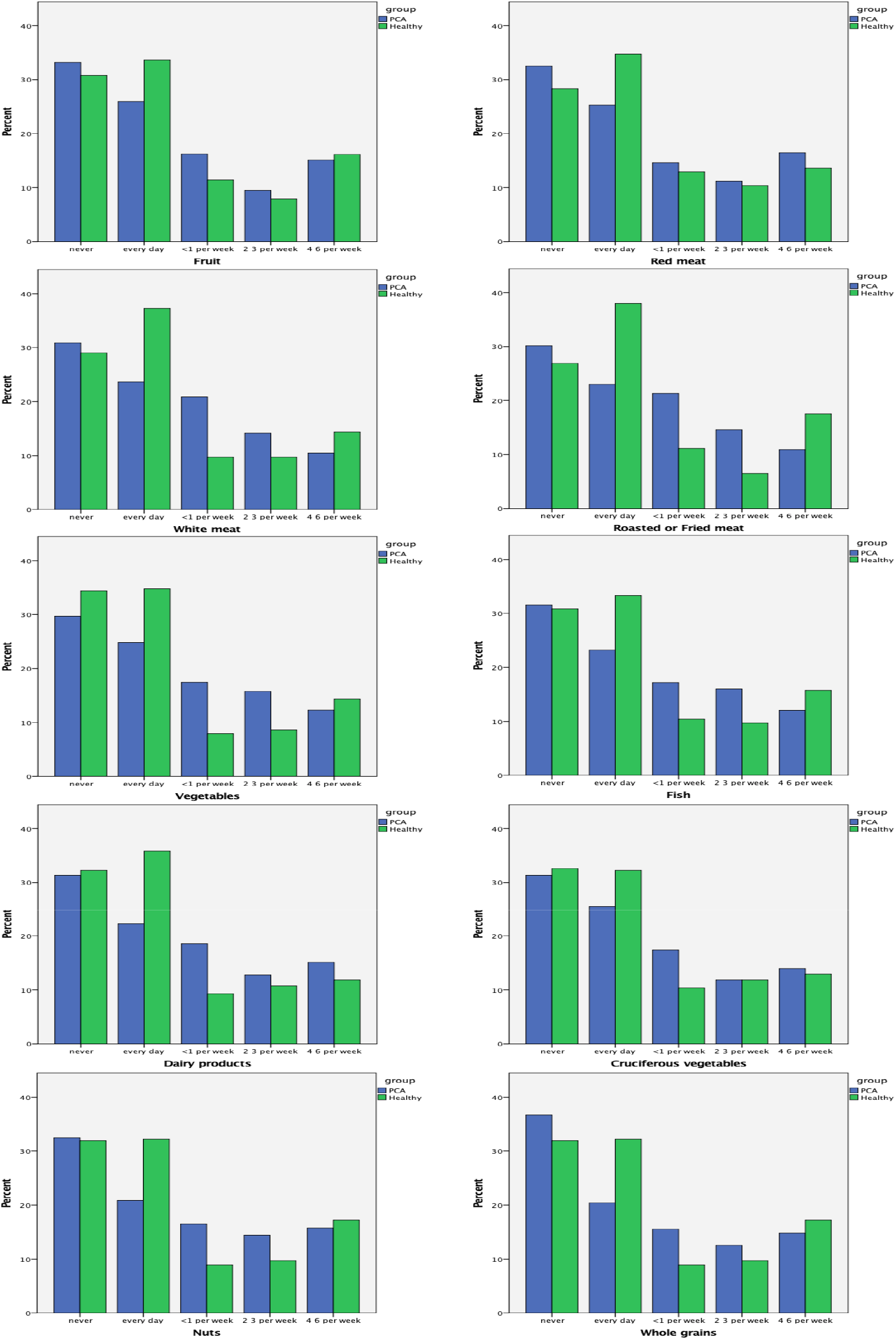
Dietary Frequencies

**Table 3:** Odds Ratios of PCa and dietary intake frequencies

|  |  |  |
| --- | --- | --- |
| **White meat** | Every day | 0.59(0.4-0.88) |
|  | <1/week | 2.03(1.2-3.38) |
| **Roasted or Fried meat** | Every day | 0.54(0.36-0.8) |
|  | 4-6/ week | 0.55(0.33-0.9) |
|  | 2-3/ week | 2.01(1.11-3.66) |
|  | <1/week | 1.71(1.04-2.81) |
| **Vegetables** | 2-3/ week | 2.12(1.24-3.63) |
|  | <1/week | 2.55(1.48-4.4) |
| **Dairy products** | Every day | 0.64(0.43-0.94) |
|  | <1/week | 2.05(1.22-3.43) |
| **Nuts** | Every day | 0.63(0.42-0.94) |
|  | <1/week | 1.8(1.06-3.06) |
| **Whole grains** | Every day | 0.55(0.37-0.81) |

Likewise, the pattern of red meat consumption was similar between the healthy and PCa groups (Chi square p=0.110). Sub-analyses showed that the rates of individuals who never, or occasionally consumed red meat did not differ significantly between the two study groups. A significantly (p=0.006) higher percentage (34.7%) of healthy men followed a daily intake of red meat.

Furthermore, differences in the intake of white meat were identified (Chi square p<0.001). Healthy men had a significantly higher tendency of consuming in a daily basis white meat (37.2% vs 23.6%, p<0.001). Although no intake frequency difference was confirmed in patients who did not introduce white meat in their diet (30.8% vs 29%), a higher rate of PCa patients (9.6% vs 20.8%, p<0.001) consumed white meat in a lower rate (1 time per week). Consumption of white meat in this frequency was associated with a high PCa incidence (Table 2, OR:2.03, 95%CI: 1.2-3.38).

A significant pooled result concerning the processed meat consumption was estimated (Chi square p<0.001). Although a higher rate of PCa patients was recorded in the <1 time per week and the 2-3 times per week comparisons (p<0.001 and p<0.001, respectively), more healthy individuals consumed processed meat in higher frequencies (4-6 times per week p=0.01, daily p<0.001). Increasing the intake frequency of processed meat, significantly reduced the PCa probability (ORs daily: 0.54, 4-6/week: 0.55, 2-3/week: 2.01, <1/week: 1.71).

Vegetable intake was significantly lower in the healthy subgroup in low consumption frequencies (<1 per week p<0.001, 2-3 per week p=0.05). Furthermore, a daily intake of vegetables was significantly higher in patients who were not diagnosed with PCa (34.7% vs 24.8% p=0.004). Therefore, an important association between dietary habits and PCa was identified (Chi square p<0.001). Infrequent consumption of vegetables was linked with twice as high PCa rate (ORs 2-3/week: 2.12, <1/week: 2.55)In addition, a higher percentage of PCa patients consumed occasionally fish (<1 per week: 17.1% vs 10.4% p=0.01, 2-3 time/week: 16% vs 9.6% p=0.01). Daily fish consumption was associated with a lower PCa rate (23.2% vs 33.3% p=0.003). Subsequently fish intake rate was significantly correlated with PCa incidence (p=0.001).

Moreover, a significant association was recorded regarding the dairy products consumption (p<0.001). PCa patients displayed a lower rate (22.2% vs 35.8% p<0.001) in daily intake and a higher percentage (18.5% vs 9.3% p<0.001) in infrequent consumption of dairy products. Daily consumption of milk and the derived products, reduced PCa rate (OR: 0.64, 95%CI: 0.43-0.94).

Cruciferous vegetables were not correlated with PCa incidence rate (p=0.07). However, fewer healthy individuals (17.4% vs 10.4% p=0.01) consumed cruciferous vegetables in a rate <1 day per week.

The daily intake (33.6% vs 20.8% p<0.001) of nuts products was higher in healthy individuals, when compared to PCa patients. In addition, infrequent nuts consumption was higher in who were diagnosed with PCa patients (<1 time per week: 16.4% vs 7.5% p<0.001). Thus, a significant correlation between nuts consumption and PCa incidence was established (p=0.001). Intake of nuts at a <1/week frequency, was linked to increased PCa prevalence (OR: 1.8, 95%CI: 1.06-3.06).

Finally, a similar pattern was identified in the intake of whole grains (p=0.001). A higher rate of PCa patients (15.5% vs 8.9% p=0.01) consumed infrequently whole grains, whereas daily intake was associated with a lower PCa incidence (20.4% vs 32.2% p<0.001). Daily consumption of whole grains displayed a significantly lower PCa OR (0.55, 95%CI: 0.37-0.81).

## DISCUSSION

An increasing body of evidence currently link PCa with dietary habits, especially in countries where a Western type of diet is consolidated. This type of diet is characterized by high rates of red or processed meat and increased amount of fat, whereas the intake of fruits, vegetables and whole grains is decreased. Another negative key feature is the high omega6:omega3 ratio and the reduced antioxidants and vitamins concentrations (15).

Mediterranean diet is a fundamental cultural component of the south European countries. It is of note that in these areas, demographic records display reduced PCa incidence and mortality rates, when compared to their northern counterparts. This was merely attributed to the increased consumption of fruits, vegetables, grains, fish, olive oil and the reduced intake of dairy products and saturated fats, main ingredients of the Mediterranean diet. These dietary products consist of naturally bioactive complexes with anti-oxidant, anti-inflammatory and alkalizing properties, which are responsible for their PCa protective effect (16).

Fruits and vegetables act as a source of microelements, carotenoids, vitamins, flavonoids and fibers (17, 18). Vegetables provide their malignancy protective role through binding and diluting carcinogens, but also by modifying specific hormonic pathways (17). Moreover, allium vegetables incorporate sulfurous phytochemicals that enhance the immune response, inhibit cell proliferation, induce apoptosis and alter the activation of androgen-responsive genes (18). Several studies confirmed the preventive effect of vegetables in PCa. Systematic cruciferous intake was, also, associated with reduced risk of PCa progression (19, 20). A meta-analysis by Liu et al. showed a significantly decrease for PCa when cruciferous vegetables were consumed (12). However, these results were not validated by Meng et al. (21). Similarly, Bosetti et al. reported a not significant correlation between cruciferous vegetables and PCa (22). In our study low volume consumption of vegetables was higher in patients who were diagnosed with PCa,, whereas healthy patients consumed vegetables on a daily basis. Noteworthy that, a non-significant association of PCa with fruits and cruciferous vegetables was established.

Red meat, processed meat and saturated fats on the other hand may enhance oncogenesis through their increased concertation of highly mutagenic carcinogens (17, 18). Apart from DNA hypomethylation, these dietary products contain heme iron, resulting in the formation of hydroxyl radicals which disrupt the signal transduction pathways (17). Although current literature suggests an increased PCa risk due to the consumption of red or processed meat, evidence is still weak (23). Even though case control studies have highlighted the harmful effect of these dietary products, a meta-analysis by Alexander et al., did not support a positive association between the intake of processed meat and PCa (24). A recent pooled analysis of 52683 PCa cases, confirmed these results (25). A correlation between frequency of red meat consumption and PCa was not justified in our study. On the contrary, there was higher consumption of processed meat in the healthy arm.

Poultry and fish intake, however, exhibit an inverse correlation with the incidence of PCa (18). Wu et al., supported that poultry intake, showed a negative association with advanced PCa (25), but a meta-analysis of 27 studies suggested that there is no linkage between white meat and PCa risk (26). Similar results were reported by Lovegrove et al., regarding fish consumption. High fish and fish oil intake did not affect the rates of PCa incidence, high grade malignancies or overall mortality (27). In our study the healthy participants tended to consume poultry and fish at higher frequencies.

Data considering milk and dairy products are contradicting (18). Mandair et al. (28), suggested an increased risk of PCa with milk consumption and Petersson et al. (29) confirmed an increased risk of disease progression in high whole milk intake. Downer et al., failed to correlate high fat milk intake with PCa disease specific mortality and a low-fat milk consumption resulted in a borderline decline of PCa patients’ death rate (30). Pooled results, however, suggest a positive interaction between dairy products and PCa risk, regardless of their fat concentration (31). Our results showed that PCa patients consumed dairy products in lower rates.

A protective effect of nuts and whole grains consumption has been extensively reported. In a retrospective analysis of 6810 PCa cases, Wang et al., found that consumption of nuts, at least five times per week, significantly reduced overall and disease specific mortality (31). Pascual-Geler et al., confirmed a statistically significant protective effect of nutritional habits containing nuts (32). Considering whole grains and fiber intake, an inverse correlation with advanced PCa incidence has been reported (33). Biochemical studies displayed a significant correlation of carcinogenesis related factors, such as tumor nuclear factor-receptor 2 (TNF-R2), e-selectin, endostatin intercellular adhesion molecule-1 (ICAM-1), vascular cell adhesion molecule-1 (VCAM-1), interleukin-6 (IL-6) and interleukin-1 receptor antagonist (IL-1RA) with the increased consumption of whole grains. As already mentioned, our study supported a higher consumption rate in healthy males.

Study limitation is the retrospective design, with no randomization and no matching between the two groups regarding demographics. Our analysis did not include any other prognostic factor for PCa, besides the dietary pattern. Considering survival analysis, a possible bias could be the lack of participants allocation in terms of disease stage and surgical and adjuvant treatment. Finally, although mMD is a valid questionnaire for the quantification of the Mediterranean dietary consumption, it is primarily self-dependent, leading to ambiguous assumptions.

## CONCLUSION

Our study showed that patients with PCa did not consume Mediterranean dietary products on a regular basis, contrary to the healthy individuals. Moreover, although, a correlation between the Mediterranean diet and the PCa incidence was not established in our series, sub-analyses displayed a higher daily intake rate of white meat, dairy products, nuts and whole grains. Therefore, we can safely conclude that an association between the Mediterranean diet and PCa should be considered. However, further trials with a larger sample size and a randomization algorithm are required to elucidate the exact effect of Mediterranean diet in the incidence and mortality of PCa patients.

## Data Availability

The data are available by the last author

## References

1. Center MM, Jemal A, Lortet-Tieulent J, Ward E, Ferlay J, Brawley O, et al. International variation in prostate cancer incidence and mortality rates. Eur Urol. 2012;61(6):1079–92.

2. Ferlay J, Shin HR, Bray F, Forman D, Mathers C, Parkin DM. Estimates of worldwide burden of cancer in 2008: GLOBOCAN 2008. Int J Cancer. 2010;127(12):2893–917.

3. Martin NE, Mucci LA, Loda M, Depinho RA. Prognostic determinants in prostate cancer. Cancer J. 2011;17(6):429–37.

4. Schwingshackl L, Hoffmann G. Does a Mediterranean-Type Diet Reduce Cancer Risk? Curr Nutr Rep. 2016;5:9–17.

5. Guasch-Ferre M, Bullo M, Martinez-Gonzalez MA, Ros E, Corella D, Estruch R, et al. Frequency of nut consumption and mortality risk in the PREDIMED nutrition intervention trial. BMC Med. 2013;11:164.

6. Schwingshackl L, Hoffmann G. Adherence to Mediterranean diet and risk of cancer: an updated systematic review and meta-analysis of observational studies. Cancer Med. 2015;4(12):1933–47.

7. Trichopoulou A, Lagiou P, Kuper H, Trichopoulos D. Cancer and Mediterranean dietary traditions. Cancer Epidemiol Biomarkers Prev. 2000;9(9):869-73.

8. Kenfield SA, DuPre N, Richman EL, Stampfer MJ, Chan JM, Giovannucci EL. Mediterranean diet and prostate cancer risk and mortality in the Health Professionals Follow-up Study. Eur Urol. 2014;65(5):887–94.

9. Bosire C, Stampfer MJ, Subar AF, Park Y, Kirkpatrick SI, Chiuve SE, et al. Index-based dietary patterns and the risk of prostate cancer in the NIH-AARP diet and health study. Am J Epidemiol. 2013;177(6):504–13.

10. Tzonou A, Signorello LB, Lagiou P, Wuu J, Trichopoulos D, Trichopoulou A. Diet and cancer of the prostate: a case-control study in Greece. Int J Cancer. 1999;80(5):704–8.

11. Hodge AM, English DR, McCredie MR, Severi G, Boyle P, Hopper JL, et al. Foods, nutrients and prostate cancer. Cancer Causes Control. 2004;15(1):11–20.

12. Liu B, Mao Q, Cao M, Xie L. Cruciferous vegetables intake and risk of prostate cancer: a meta-analysis. Int J Urol. 2012;19(2):134–41.

13. Karatzas A, Giannatou E, Tzortzis V, Gravas S, Aravantinos E, Moutzouris G, et al. Genetic polymorphisms in the UDP-glucuronosyltransferase 1A1 (UGT1A1) gene and prostate cancer risk in Caucasian men. Cancer Epidemiol. 2010;34(3):345–9.

14. von Elm E, Altman DG, Egger M, Pocock SJ, Gotzsche PC, Vandenbroucke JP, et al. The Strengthening the Reporting of Observational Studies in Epidemiology (STROBE) statement: guidelines for reporting observational studies. J Clin Epidemiol. 2008;61(4):344–9.

15. Chua ME, Sio MC, Sorongon MC, Dy JS. Relationship of dietary intake of omega-3 and omega-6 Fatty acids with risk of prostate cancer development: a meta-analysis of prospective studies and review of literature. Prostate Cancer. 2012;2012:826254.

16. Lopez-Guarnido O, Alvarez-Cubero MJ, Saiz M, Lozano D, Rodrigo L, Pascual M, et al. Mediterranean diet adherence and prostate cancer risk. Nutr Hosp. 2014;31(3):1012–9.

17. Kruk J, Aboul-Enein H. What Are the Links of Prostate Cancer with Physical Activity and Nutrition?: A Systematic Review Article. Iran J Public Health. 2016;45(12):1558–67.

18. Lin PH, Aronson W, Freedland SJ. Nutrition, dietary interventions and prostate cancer: the latest evidence. BMC Med. 2015;13:3.

19. Hsing AW, Chokkalingam AP, Gao YT, Madigan MP, Deng J, Gridley G, et al. Allium vegetables and risk of prostate cancer: a population-based study. J Natl Cancer Inst. 2002;94(21):1648–51.

20. Richman EL, Carroll PR, Chan JM. Vegetable and fruit intake after diagnosis and risk of prostate cancer progression. Int J Cancer. 2012;131(1):201–10.

21. Meng H, Hu W, Chen Z, Shen Y. Fruit and vegetable intake and prostate cancer risk: a meta-analysis. Asia Pac J Clin Oncol. 2014;10(2):133–40.

22. Bosetti C, Filomeno M, Riso P, Polesel J, Levi F, Talamini R, et al. Cruciferous vegetables and cancer risk in a network of case-control studies. Ann Oncol. 2012;23(8):2198–203.

23. Ma RW, Chapman K. A systematic review of the effect of diet in prostate cancer prevention and treatment. J Hum Nutr Diet. 2009;22(3): 187-99; quiz 200-2.

24. Alexander DD, Mink PJ, Cushing CA, Sceurman B. A review and meta-analysis of prospective studies of red and processed meat intake and prostate cancer. Nutr J. 2010;9:50.

25. Wu K, Spiegelman D, Hou T, Albanes D, Allen NE, Berndt SI, et al. Associations between unprocessed red and processed meat, poultry, seafood and egg intake and the risk of prostate cancer: A pooled analysis of 15 prospective cohort studies. Int J Cancer. 2016;138(10):2368–82.

26. He Q, Wan ZC, Xu XB, Wu J, Xiong GL. Poultry consumption and prostate cancer risk: a meta-analysis. PeerJ. 2016;4:e1646.

27. Lovegrove C, Ahmed K, Challacombe B, Khan MS, Popert R, Dasgupta P. Systematic review of prostate cancer risk and association with consumption of fish and fish-oils: analysis of 495,321 participants. Int J Clin Pract. 2015;69(1):87–105.

28. Mandair D, Rossi RE, Pericleous M, Whyand T, Caplin ME. Prostate cancer and the influence of dietary factors and supplements: a systematic review. Nutr Metab (Lond). 2014;11:30.

29. Pettersson A, Kasperzyk JL, Kenfield SA, Richman EL, Chan JM, Willett WC, et al. Milk and dairy consumption among men with prostate cancer and risk of metastases and prostate cancer death. Cancer Epidemiol Biomarkers Prev. 2012;21(3):428–36.

30. Downer MK, Batista JL, Mucci LA, Stampfer MJ, Epstein MM, Hakansson N, et al. Dairy intake in relation to prostate cancer survival. Int J Cancer. 2017;140(9):2060–9.

31. Aune D, Navarro Rosenblatt DA, Chan DS, Vieira AR, Vieira R, Greenwood DC, et al. Dairy products, calcium, and prostate cancer risk: a systematic review and meta-analysis of cohort studies. Am J Clin Nutr. 2015;101(1):87–117.

32. Pascual-Geler M, Urquiza-Salvat N, Cozar JM, Robles-Fernandez I, Rivas A, Martinez-Gonzalez LJ, et al. The influence of nutritional factors on prostate cancer incidence and aggressiveness. Aging Male. 2018;21(1):31–9.

33. Tabung F, Steck SE, Su LJ, Mohler JL, Fontham ET, Bensen JT, et al. Intake of grains and dietary fiber and prostate cancer aggressiveness by race. Prostate Cancer. 2012;2012:323296.

